# Developing and comparing machine learning models to detect sleep apnoea using single-lead electrocardiogram (ECG) monitoring

**DOI:** 10.1101/2021.04.19.21255733

**Authors:** Marcel Hedman, Alex Rojas, Anmol Arora, David Ola

**Affiliations:** Harvard University, United States; Nural Research, United Kingdom; School of Clinical Medicine, University of Cambridge, United Kingdom

**Keywords:** machine learning, artificial intelligence, sleep medicine, polysomnography, telemonitoring, remote patient monitoring, telemedicine

## Abstract

**Background:** Sleep apnoea has a high disease burden but remains underdiagnosed, in part due to the expensive and resource intensive nature of polysomnography, its definitive investigation. Emerging literature suggests that it may be possible to detect sleep apnoea using single-lead ECG signals, such as those obtained from smartwatches. In this study, we use two forms of recurrent neural networks (RNNs) to detect sleep apnoea events from single-lead ECG signals.

**Methods:** We use single-lead ECG data from the PhysioNet Apnea-ECG database, which contains data from 70 patients. We train a bidirectional gated recurrent unit (GRU) model and a bidirectional long short-term memory (LSTM) model on labelled ECG signals from 35 patients and test the models on the remaining 35 patients in the dataset.

**Results:** Both models achieved 97.1% accuracy, sensitivity and specificity to detect whether the ECG recordings belonged to a patient diagnosed with sleep apnoea. This corresponds to 34/35 patients in the dataset. At detecting individual apnoea events, the GRU and LSTM models achieved 90.4% and 91.7% accuracies respectively.

**Discussion:** The models achieved high levels of accuracy, specificity and sensitivity. Bidirectional RNNs are strengthened by the ability of the models to be informed by both past and future states when analysing sequential data, such as ECGs. The models also require minimal human intervention as they automatically extract features from the data. If single-lead ECGs prove a suitable tool for sleep apnoea detection, this may enhance the diagnosis of sleep apnoea and potentially allow widespread screening for the condition.

**Conclusions:** We note that using models such as bidirectional RNNs has the potential to augment model performance. However, more research and validation is required in order to test whether these may be applicable to other datasets and in clinical practice.

## Introduction

Sleep Apnoea (SA) is characterised by intermittent interruptions in airflow during sleep [1]. Sleep apnoea can be categorised into three types: 1. central sleep apnoea, which can be triggered by the brain failing to activate breathing muscles, 2. obstructive sleep apnoea, the collapse of the airway during sleep, or 3. a combination of both [2]. The incidence of sleep apnoea has been estimated to be as high as 20% of the population and it has been suggested that approximately 90% of cases may be undiagnosed [3] [4].

While asleep, one apnoea event is defined as a >90% reduction in airflow lasting at least 10 seconds. A hypopnoea is a less severe reduction in airflow. The severity of sleep apnoea is graded by the number of apnoea and hypopnoea events per hour, the Apnoea Hypopnoea Index (AHI), with more than 5 events per hour being diagnostic [5]. The definitive method for diagnosing for sleep apnoea is polysomnography, which is resource intensive and complex to interpret. A polysomnogram consists of several simultaneous measurements: e.g. electroencephalogram (EEG), electrooculogram (EOG), electromyogram (EMG), electrocardiogram (ECG), pulse oximetry and nasal and oral airflow. In recent years, there has been development of home sleep apnoea tests which allow monitoring of patients at home rather than in a controlled clinical setting but these are still resource intensive and involve monitoring several parameters simultaneously. The limited availability of centres and clinicians able to diagnose sleep apnoea leads to long waiting lists for the patients seeking a diagnosis [6].

Electrocardiogram (ECG) is a test performed to monitor the electrical activity of the heart. ECGs can be performed with varying numbers of leads, typically ranging from one to twelve. In this study, we train machine learning models on data from single-lead ECG investigations to identify sleep apnoea. We use the Physionet Apnea-ECG Database which consists of single-lead ECG signals from 70 patients, 35 allocated for training a model and 35 allocated for its testing [7]. There have been efforts to analyse single-lead ECG signals previously, which have yielded respectable levels of accuracy, specificity and sensitivity. The machine learning models previously tested include feature engineering and feature learning methods. Feature engineering models require processing of raw data to extract features which may be useful in analysing unseen data. Feature learning models are designed to learn the features themselves, reducing the need for pre-processing. Examples of feature learning methods which have been applied to the Physionet database include Singh and Majumder’s Pre-trained AlexNet CNN + Decision Fusion model and Wang et al’s LeNet-5 CNN model [2] [8]. Examples of feature engineering based methods include: Sharma and Sharma’s Feature Engineering + LS-SVM and Song et al’s Feature Engineering + HMM-SVM [9] [10].

In this study, we develop novel models using recurrent neural network (RNN) variants such as gated recurrent unit (GRU) and long short-term memory (LSTM). The models we prepared are feature learning models with some pre-processing of data to identify R-R peaks and amplitudes from the ECG signals. We use the PhysioNet Apnea-ECG dataset pre-processing scheme provided by Wang et al [2]. If the analysis of single-lead ECG is proved to be an accurate method of diagnosing sleep apnoea, there is potential to simplify and broaden the screening for sleep apnoea diagnosis given that single-lead ECG data are being made increasingly accessible through 21st century smart watches.

## Methods

### Sleep apnoea dataset

In this study, we use the PhysioNet Apnea-ECG dataset provided by Philipps University [7]. The dataset consists of 70 records, divided into a learning set and a testing set of 35 records each. The ECG data were sampled at 100Hz with durations ranging between 401 and 587 min. Data are labelled once per minute in each ECG signal, indicating whether or not an apnoea event was in progress at the beginning of that minute. Both apnoea and hypopnoea events were simply labelled as apnoea events. The dataset does include other signals, including chest and abdominal respiratory effort, oronasal airflow and oxygen saturation, however in this study only ECG signals were used for analysis.

### Pre-processing of data

We processed our data in line with the method outlined by Wang et al [2]. This involved extracting the R-R intervals and amplitudes from single-lead ECG using the Hamilton algorithm plus the positions and values of calculated R-peaks.

An ECG tracks the electrical activity of the heart, with each beat forming a QRS complex. The QRS complex incorporates the Q, R, and S waves, which represents ventricular depolarisation [11]. R-peaks are crucial for analysing this QRS complex [12], and using the aforementioned pre-processing scheme we can extract RR-intervals and amplitudes. This implementation takes into account the adjacent two segments of each labelled segment because adjacent segment information has been show to be helpful for per-segment sleep apnoea detection [2].

### Machine Learning Models

The input of the models is the R-R intervals and amplitudes from the single-lead ECG signal and the output is whether or not the input patient has sleep apnoea. It is extremely important to attain models with high accuracy that are not overparametrized [13]. In conjunction with our goal of deployment, we seek to maximize accuracy while constraining model size during training such that these models can be efficient.

### Model 1: LSTM with Convolution

The model architecture involves both convolution and recurrent neural network properties; the input is first passed to a single convolution layer filter size of 5 and stride 2, and the activation map is then passed to max-pooling which subsamples by a factor of 3. Next, we repeat twice bidirectional layers with 64 LSTM nodes, followed by dropout with *p* = .8 and batch normalization. Finally, we pass through fully connected layers with ReLU activation and finally an output layer with softmax activation. This model thus has 760,258 parameters and was trained with an Adam optimizer with a learning rate of 5 ·10^−3^, with a learning rate scheduler that decreased the learning rate after 70 epochs by 1 ·10^−1^ every 10 epochs. In total, we trained for 100 epochs and built the model with Tensorflow.

### Model 2: GRU with Convolution

This model is almost identical to Model 1, implementing recurrence on top of convolution. Yet, we replaced the LSTM nodes with GRU nodes. This model was trained identically to Model 1, and slightly cuts down on the number of parameters with 723,906.

## Results

We evaluated the performance of our models at both a per minute (segment) apnoea detection level as well as on a per patient (recording) level. This is in line with the traditional method of determining whether a person is diagnosed with sleep apneoa based on the frequency of apnoea events. We use five metrics to evaluate the machine learning model:

- Accuracy: Accuracy measures the percentage of predicted labels that matches the ground truth labels as defined by the creators of the dataset.
- Specificity: Specificity measures the true negative rate, this is the percentage of true negative predictions over the total number of predictions of the negative class.

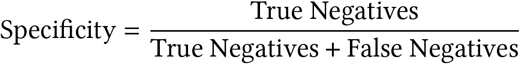
- Sensitivity: Measures the true positive rate, which is the percentage of all positive predictions which are true positives.

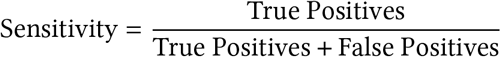
- Area under receiver operating characteristic (ROC) curve (AUC): The ROC curve shows the balance of true positive rate to false positive rate as the threshold boundary for classification to a binary class is changed. The area under it reaches a maximum of 1.
- Correlation: The correlation measures the strength of the relationship between two variables and was used to compare the predicted AHI index to the actual index. This is in line with previous studies [2] [9].

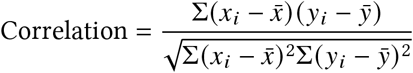

### Per Segment

Table 1 contains the results across our approaches and it can be seen that both the LSTM and GRU models outperform a previously constructed modified LeNet5 model which we use as a benchmark [2].

**Table 1:**
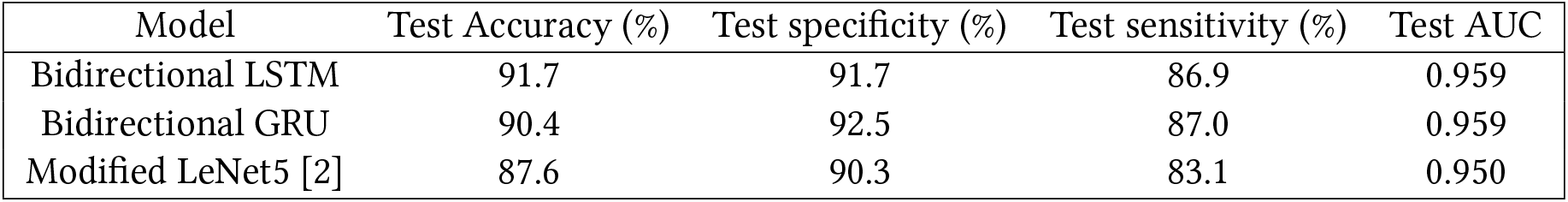
Per segment results of the bidirectional RNN models

| Model | Test Accuracy (%) | Test specificity (%) | Test sensitivity (%) | Test AUC |
| --- | --- | --- | --- | --- |
| Bidirectional LSTM | 91.7 | 91.7 | 86.9 | 0.959 |
| Bidirectional GRU | 90.4 | 92.5 | 87.0 | 0.959 |
| Modified LeNet5 [2] | 87.6 | 90.3 | 83.1 | 0.950 |

Figure 1 illustrates the receiver operating characteristic (ROC) curve for the high-performing bidirectional GRU model. The AUC is calculated as 0.959, which represents outstanding model performance [14].

**Figure 1:**
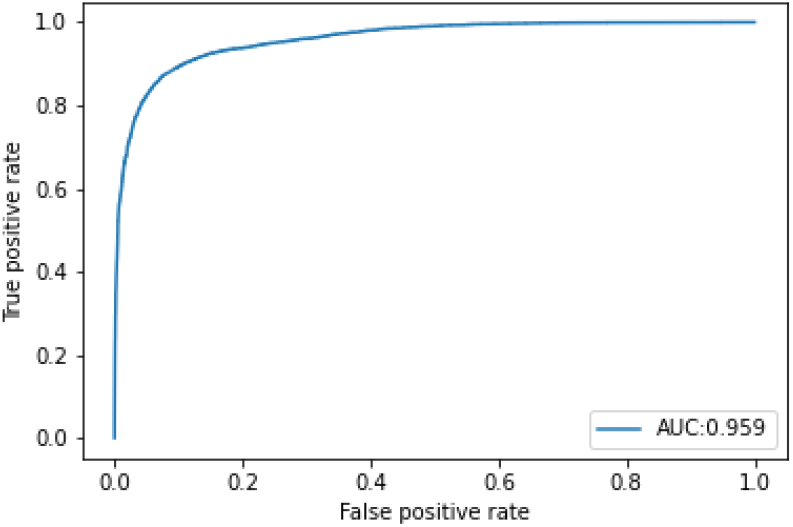
Bidirectional GRU ROC curve

### Per Patient

Given the ability to classify individual apnoea events in sleep, a classification then needs to be made on a per patient level. To perform this classification, we used the AHI score. The AHI score gives the number of apnoea and hypoapnoea events per hour and scores less than 5 in an asymptomatic patient are considered normal [15]. Within this dataset, there was an intermediate class for borderline cases and when this dataset was used in a competition, it was advised that these borderline cases need not be used for testing the model [16]. However, in order to ensure that our models correspond with currently accepted clinical practice, we used a simple cut-off of an AHI of 5 when determining the diagnosis. Both models accurately classified 34/35 patients as having sleep apnoea or not, based on the AHI definition. The one patient which the algorithms incorrectly classified was one of the aforementioned borderline cases.

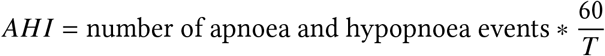

Table 2 demonstrates that the LSTM and GRU models had high performance in determining whether a recording belonged to someone diagnosed with sleep apnoea and this is to be expected given the high performance on the per segment predictions. The models, therefore, may be potentially highly effective at screening sleep apnoea.

**Table 2:** Per recording results

| Model | Test Accuracy (%) | Test specificity (%) | Test sensitivity (%) | Corr |
| --- | --- | --- | --- | --- |
| Bidirectional LSTM | 97.1 | 91.7 | 100.0 | 0.966 |
| Bidirectional GRU | 97.1 | 100.0 | 95.7 | 0.969 |
| Modified LeNet5 [2] | 97.1 | 91.7 | 100.0 | 0.943 |

## Discussion

### Summary of Key Findings

In this study, we developed and tested two models to detect sleep apnoea using the Physionet Apnea-ECG dataset. We found that both the bidirectional GRU and LSTM models achieved 97.1% accuracy, with both high sensitivity and specificity, in predicting whether a patient would have sleep apnoea.

### Comparison to Existing Literature

Even though sleep apnoea is usually a respiratory disease, it is well recognised that it is associated with cardiac rhythm disturbances which may be detected by ECG [17] [18]. The use of ECG alone is not routinely used to diagnose sleep apnoea. However, the use of a two-lead ECG does form part of the polysomnogram which is the current gold-standard method of diagnosing sleep apnoea. The ECG is recognised to include a range of information which is becoming increasingly interpretable with the advent of machine learning analysis [19].

Previous studies attempting to diagnose sleep apnoea with single-lead ECGs or other parameters such as SpO2 have used several different types of models, including deep vanilla neural networks (DVNNs), artificial neural networks (ANNs), convolutional neural networks (CNNs) and recurrent neural networks (RNNs) [6] [20]. Table 3 compares the results of models generated in this study to models produced in several selected preceding studies. A systematic review in 2019 has provided a more thorough analysis of previous attempts to diagnose sleep apnoea using single-lead ECGs, with the models in this study achieving higher levels of accuracy than most of those identified in the review [6]. However, it should be noted that this high accuracy in a small dataset may not necessarily translate to being the most clinically useful.

**Table 3:** Per recording performance comparison of our models with selected models using the same Physionet Apnea-ECG dataset

| Author | Dataset | Model | Accuracy (%) | Specificity (%) | Sensitivity (%) | Correlation |
| --- | --- | --- | --- | --- | --- | --- |
| Our study | Apnea-ECG | Bidirectional LSTM | 97.1 | 91.7 | 100.0 | 0.966 |
| Our study | Apnea-ECG | Bidirectional GRU | 97.1 | 100.0 | 95.7 | 0.969 |
| Wang et al (2019) [2] | Apnea-ECG | Modified LeNet5 | 97.1 | 91.7 | 100.0 | 0.943 |
| Li et al (2018) [31] | Apnea-ECG | Decision Fusion | 100.0 | 100.0 | 100.0 | - |
| Sharma and Sharma (2016) [9] | Apnea-ECG | LS-SVM | 97.1 | 100.0 | 95.8 | 0.841 |
| Song et al (2016) [10] | Apnea-ECG | HMM-SVM | 97.1 | 100.0 | 95.8 | 0.860 |
| Chang et al (2020) [32] | Apnea-ECG | 1D deep CNN | 97.1 | 100.0 | 95.7 | 0.865 |

### Comparison of Machine Learning Models

#### Artificial Neural Networks

Artificial neural networks (ANN) are computational models designed as a simplified model of the human brain, consisting of layers of nodes and non-linear activation that can model complex functions. Multiple layers form a deep network, and these models train by minimizing a loss function through backpropagation and gradient-descent [21]. ANNs have been demonstrated to predict heartbeat classification tasks with high degrees of accuracy for R-R intervals of ECG recordings [22]. However, these models become overfit when trained on small datasets, failing to generalize well. This overfitting problem is often addressed by a technique known as regularisation which stops any individual weight in the network from growing too large [23]. Within a neural network, this is achieved through dropout which randomly sets nodes to zero in training.

#### Convolutional Neural Networks

Standard ANNs suffer when applied to certain tasks including image recognition, as they fail to be translation-invariant and contain many parameters to be trained. A convolutional neural network (CNN) addresses these challenges. The CNN was first introduced as the time delay neural network, creating a translation-invariant network trained with backpropagation for the first time [24]. Originally taking inspiration from receptive fields created by neurons in the visual cortex, these networks are most common for image classification, but can be applied more broadly for feature extraction. CNNs build upon the ANN by introducing convolution, a process by which trainable filters convolve over an input and output an activated feature map, which has the major advantage of reducing the number of trainable parameters for a similar sized network. A common activation function introduced to CNN by AlexNet, a CNN trained on the ImageNet database, is the non-linear ReLU [25]. After the convolutional layers, the activation maps are flattened and passed into a standard ANN. Max-pooling is a method for downsampling the activation maps between certain layers designed to reduce dimensionality by selecting the maximally activated convolved node and discarding the rest.

#### Recurrent Neural Networks

One weakness of the ANN and CNN is that they can not process data that has inherent sequential meaning in a way that retains memory. The recurrent neural network (RNN) addresses this problem by passing through in each forward pass a hidden state *h*_*t*_, which taken with the input allows the network to retain memory about the past, often for sequential tasks such as processing language. This is updated by passing the current state and hidden state through an activation function after an affine transformation. However, these networks are subject to the vanishing gradient problem, and to address this, two architectures exist: the Gated Recurrent Unit (GRU) and the Long-Short Term Memory Unit (LSTM) networks.

One earlier attempt to address this is the LSTM, which used additional affine transformations and element-wise selection on the hidden state and inputs to update the states and activated state [26]. The LSTM hidden state is computed where the output gate *o*_*t*_, an activated affine transformation of the input, previous hidden state, and *c*_*t*_ moderates how much of the activated *c*_*t*_ to pass through. We have that *c*_*t*_ is also computed as an activated affine transformation of the input and previous hidden state with a forget gate and input gate. We can introduce bidirectionality, which allows the network to learn from both the future inputs and past inputs [27]. LSTMs have been shown to perform on ECG data generally and this is to be expected due to its memory ability, it was therefore a natural extension to apply it to sleep apnoea detection using single-lead ECG [28]. A simplification of the LSTM came in the form of the GRU [29].

The two bidirectional models we produced have the advantage that they can learn from both past (backwards) and future (forwards) states, which is a useful characteristic when analysing data with long-term dependencies between time steps in sequential data [30]. RNNs are ideally suited to time-series data such as ECGs, as applied in this study. The models used in this study are classified as ‘feature learning’, such that they require no manual feature engineering. In our case, human processing the data was limited to pre-processing of the data to extract R-R peaks and amplitudes.

### Limitations

In this dataset, hypopnoea events were labelled as apnoea events which means that our models are unable to distinguish between the two [33]. Furthermore, the dataset only contains apnoeas which are obstructive or mixed, with no episodes of purely central apnoea. Future iterations of these models should involve larger datasets which are labelled for both apnoea and hypopnoea events, although both are used when calculating AHI. The small sample size used in this study dictates that further research is required to validate these findings. Only one dataset was used, in which the same ECG techniques were applied. In order for these findings to have implications for future practice, the machine learning system must be interoperable. Without testing a more diverse dataset, we are unable to comment on whether our model would be effective if applied to novel datasets or data taken from other ECG monitoring devices. The purpose of this study was to highlight potential models which offer most potential for future avenues of research. Such future avenues should involve external validation and randomised trials conforming to SPIRIT-AI and CONSORT-AI guidelines in order to truly determine whether the algorithms may be suitable for clinical practice [34] [35].

It should be noted that in clinical practice, sleep apnoea diagnosis does not solely rely on AHI but also on the clinical presentation and symptoms experienced by the patient [16]. Sleep studies are to be used in combination with other clinical tools, for example the Epworth Sleepiness Scale [36].

In recent years, the development of machine learning algorithms has led to the issue of ‘black-box’ algorithms, whereby it is difficult to understand how the outputs are derived. In this case, the machine learning models were able to detect whether a single-lead ECG signal indicated sleep apnoea, which is a difficult task for even the most skilled ECG readers. In current clinical practice, diagnoses of sleep apnoea are derived from polysomnograms, which is both an expensive and resource intensive method. Conversely, it is also difficult to monitor sleep apnoea at home, given that the resources required are typically unavailable outside a secondary care setting.

### Implications of Findings

In recent years, there has been an exponential increase in the adoption of wearable devices, including Apple watches, Oura rings and other smartwatches. This has led to the use of such devices to assist in remote patient monitoring. Certain novel watches are capable of providing ECG monitoring as well as measuring breathing rate in some cases. The use of ECG watches to predict, diagnose and monitor sleep apnoea seems to be within the realms of possibility in the context of a growing body of literature suggesting that it might be possible to diagnose using only a single-lead ECG. There remain a number of uncertainties regarding how such practice would be implemented, including whether the analysis of ECG leads would occur on the device, such as in edge computing, or centrally. The issue of accountability has been raised in the literature as a key barrier to adoption of AI systems, since there is a plausible risk that the systems would have imperfect accuracy.

The potential use of AI in clinical practice is accompanied by discussion of practical and ethical issues that may surround its implementation, including the need for interpretability of algorithms [37]. Although the models were able to detect sleep apnoea with a high degree of accuracy, it is often unclear how this is achieved. Other ethical arguments relate to the interaction between the AI system and the clinicians providing care to the patient since there is a need to supervise the models, which is difficult if the operator has a limited understanding of how the models are operating [38].

There are a number of obstacles before such technologies may be used in practice. Yoo et al highlight three characteristics of a transformative innovation which are relevant in this instance: interoperability, homogeneity of data and self-referential nature [39] [40]. In order for algorithms to be useful in clinical practice, it is important to ensure that they are interoperable, in such a way that they are generalisable to different sources of input data e.g. different devices collecting ECG signals in different individuals. At the very least, there needs to be an understanding of any limitations of the generalisability of the model. There should also be effort spent trying to homogenise data between different devices, or data sources, in order to make achieving interoperability a realistic prospect. Lastly, the utility of an algorithm would increase substantially if it could be used to improve the quality of or generate other algorithms. For example, a system developed to interpret ECG signals in such a way that it can detect sleep apnoea may further be applied to detect other conditions, such as through the implementation of techniques such as transfer learning.

Unfortunately, there is a notable shortage of single-lead ECG datasets labelled for sleep apnoea events. In order to develop the findings of this study and to improve the models to a standard which would be acceptable for routine care, there is a need to develop larger datasets with more diverse populations. A number of studies assessing the utility of machine learning algorithms to diagnose sleep apnoea have used the same datasets, limiting the interoperability and generalisability of findings. This study as well as previous studies are important in proving that there is potential for machine learning models to detect sleep apnoea using only a single-lead ECG signal, however further research is required using larger datasets. The accretion of datasets for developing models is complicated by the fact that it is difficult for human clinicians to label single-lead ECG signals with sleep apnoea diagnoses without clinical context. Developing large scale datasets, which includes patients known to have sleep apnoea remains a priority for advancing research in the field.

## Conclusions

This study adds to the growing body of literature suggesting that, in the future, single-lead ECGs may offer potential to diagnose sleep apnoea using only a smartwatch. We propose two novel machine learning models applied to ECG analysis to detect sleep apnoea. Both the bidirectional GRU and the bidirectional LSTM models outperformed most models in existing literature. They highlight the potentially important role of recurrent neural networks when developing future models. Further research is required to confirm these findings using larger datasets and data obtained using smartwatches.

## Data

All data are freely accesible on Physionet [7]: https://physionet.org/content/apnea-ecg/1.0.0/

## Data Availability

All data are freely accesible on Physionet [7]: https://physionet.org/content/apnea-ecg/1.0.0/

